# Early Initiation of Fast-Track Care for Persons Living with HIV Initiating Dolutegravir-Based Regimens during a Period of Severe Civil Unrest in Port-au-Prince, Haiti

**DOI:** 10.1101/2024.08.12.24311216

**Authors:** Jean Bernard Marc, Samuel Pierre, Othnia Ducatel, Fabienne Homeus, Abigail Zion, Vanessa R. Rivera, Nancy Dorvil, Patrice Severe, Colette Guiteau, Vanessa Rouzier, Ingrid T. Katz, Carl Frederic Duchatelier, Guyrlaine Pierre Louis Forestal, Josette Jean, Guirlaine Bernadin, Emelyne Droit Dumont, Rose Cardelle B. Riche, Jean William Pape, Serena P Koenig

## Abstract

**Introduction:** Differentiated service delivery (DSD) models have been widely implemented for patients who are established in HIV care. However, DSD has rarely been offered to those newly diagnosed with HIV. We conducted a study to assess the effectiveness of early fast-track care during the COVID-19 pandemic and a period of severe civil unrest at GHESKIO, in Haiti.

**Methods:** We conducted a pilot randomized trial among adults presenting with World Health Organization Stage 1 or 2 disease at HIV diagnosis to determine whether early fast-track care (at eight to 12 weeks after same-day HIV testing and ART initiation) was associated with superior outcomes, compared with standard care (deferred eligibility for fast-track care). All participants received tenofovir disoproxil fumarate/lamivudine/dolutegravir (TLD), and HIV-1 RNA <200 copies/mL was required prior to initiating fast-track care. The primary outcome was 48-week HIV-1 RNA <200 copies/mL, with intention-to-treat analysis.

**Results:** From December 13, 2020, to August 19, 2022, 247 participants were randomized; 2 met protocol-specified criteria for late exclusions, leaving a study population of 245 (standard: 116; early fast-track: 129). All participants initiated TLD on the day of HIV diagnosis. In the standard group, 2 (1.7%) died, 106 (91.4%) were retained in care, and 78 (67.2%) received 48-week viral load testing; 66 (84.6% of those tested; 56.9% of those randomized) had HIV-1 RNA <200 copies/mL. In the early fast-track group, 1 (0.8%) died, 112 (86.8%) were retained in care, and 87 (67.4%) received 48-week viral load testing; 79 (90.8% of those tested; 61.2% of those randomized) had 48-week HIV-1 RNA <200 copies/mL. There was no difference in primary outcome (48-week HIV-1 RNA <200 copies/mL) between the early fast-track and standard groups (61.2% vs. 56.9%; RD: 0.043; 95% CI: -0.080, 0.167).

**Conclusions:** The provision of fast-track care as early as 8 weeks after TLD initiation is associated with high levels of retention in care and viral suppression in a setting of severe civil unrest, with no difference in outcome compared to deferred eligibility for fast-track care. Completion of 48-week viral load testing was suboptimal; low-cost point-of-care testing capacity may facilitate completion of viral load testing in this setting.

## INTRODUCTION

Differentiated service delivery (DSD) models have been widely implemented to facilitate follow-up care for persons living with HIV (PLWH). Multiple studies have demonstrated excellent outcomes with DSD strategies, including fewer visits, multi-month dispensing of medications, and decentralized drug distribution.^1-10^ However, expedited services are generally offered only to PLWH considered to be stable after at least 6 months of antiretroviral therapy (ART).^1,7,8^ Data on outcomes of earlier DSD eligibility are limited. With widespread use of dolutegravir (DTG)-based ART regimens in low and middle-income countries (LMICs), viral suppression can be achieved within eight to 12 weeks.^11^ This provides the potential opportunity for earlier transition to expedited services, during the period when loss to follow up (LTFU) rates are the highest.

We conducted a pilot randomized trial to determine whether early fast-track care (at eight to 12 weeks after ART initiation) was associated with superior outcomes, compared with standard care (deferred eligibility for fast-track care) in Haiti. We hypothesized that early fast-track care would improve the primary outcome of retention with viral suppression at 48 weeks after enrollment.

## METHODS

### Study Design and Setting

We conducted a pilot randomized trial to compare rates of viral suppression with early fast-track versus standard care among patients newly diagnosed with HIV at the Haitian Group for the Study of Kaposi’s Sarcoma and Opportunistic Infections (GHESKIO) in Port-au-Prince, Haiti. The adult HIV prevalence in Haiti is an estimated 1.7%.^12^ This study was conducted during the COVID-19 pandemic, and during a period of severe political instability, civil unrest, and gang violence in Haiti.^13^

### Ethics Statement

The study was approved by the institutional review boards at GHESKIO, Brigham and Women’s Hospital, and Weill Cornell Medical College. Written informed consent was obtained from all participants.

### Study Participants

Patients newly diagnosed with HIV were recruited for this study at the GHESKIO outpatient clinic. Key inclusion criteria included ≥18 years of age and WHO Stage 1 or 2 disease; exclusion criteria included previous ART, pregnancy, creatinine clearance (CrCl) <50 or alanine transaminase (ALT) or aspartate transaminase (AST) >5 times the upper limit of normal.

### Randomization and Masking

Participants were randomized in a 1:1 ratio using a computer-generated random-number list in the GHESKIO Data Management Unit. Study staff were not blinded to randomization group.

### Study Procedures

All participants initiated TLD on the day of HIV diagnosis and were enrolled and randomized within the subsequent three days. Specimens were collected for creatinine, ALT, AST, complete blood count (CBC), and CD4 count testing. Same-day lab results were not available, so participants meeting exclusion criteria were late exclusions.

The only difference between the two groups was the timing of eligibility for fast-track care, which was provided in the same manner for both groups. In the standard group, the visit schedule initially included monthly visits for the first six months, and then quarterly visits. However, in mid-2021, the standard of care changed to quarterly visits throughout the first year in care. At the 24-week visit, participants meeting eligibility criteria initiated fast-track care. In the early fast-track group, participants received the same care as the standard group for the first eight weeks. At the week 8 visit, viral load testing was conducted, and participants who met eligibility criteria initiated fast-track care; those who were not yet eligible were re-assessed at the week 12 visit.

For both groups, eligibility for fast-track care required being on time for the clinic visit (≤3 days late), without an active WHO Stage 3 or 4 condition, and with HIV-1 RNA <200 copies/mL. Fast-track care included expedited, focused visits with minimal waiting time; pre-packaged point-of-service dispensing of ART and other medications during the clinic visit; and quarterly visits at either the GHESKIO facility or one of nine community ART sites. Retention activities included phone calls prior to visits and after missed visits, a transportation subsidy of 100 Haitian gourdes (about $US 1.00) and a phone card (valued at 100 Haitian gourdes) at each visit. Participants with HIV-1 RNA ≥200 copies/mL received adherence counseling, followed by repeat viral load testing.

### Outcomes

The primary outcome was the proportion of participants with HIV-1 RNA <200 copies/mL at 48 weeks after enrollment, with a prespecified window of 47 to 60 weeks after enrollment; missing test results were considered virologic failures. Secondary outcomes included: 48-week HIV-1 RNA cut-offs of <50 and <1,000 copies/mL; adherence as measured by medication possession ratio (proportion of days ART was dispensed within the study period); and tolerability, as measured by permanently discontinuing TLD due to adverse events. Participants were considered retained in care at 48-weeks if they attended a visit from 24 to 72 weeks after enrollment. Lost to follow-up (LTFU) was defined as lost to care without known death; participants were classified as having fled their homes due to gang violence if it was reported to study staff.

### Statistical Analysis

For the primary outcome, a sample size of 242 was calculated to provide at least 80% power to detect an absolute difference of 11% in the proportion of participants with 48-week HIV-1 RNA <200 copies/mL (standard: 70%; early fast-track: 81%) assuming a significance level of 0.25, using the Chi-square test. The sample size was inflated to 247 to account for late exclusions.

We compared the proportion of participants with 48-week HIV-1 RNA <200 copies/mL (primary endpoint) and binary secondary outcomes using the Chi-square test (Fisher’s exact test was used for sparse data). We presented unadjusted risk difference (RD) with 95% confidence interval (CI) using the Wilson score interval. We compared the mean number of visits using the mean difference (MD) with 95% CI computed using a pooled T-test. All analyses were conducted using SAS version 7.1. The study was registered with ClinicalTrials.gov NCT04311944.

## RESULTS

From December 13, 2020, to August 19, 2022, 252 patients were screened and 247 (98.0%) were enrolled and randomized (**Fig 1)**. Two participants met pre-specified criteria for late exclusions, leaving a total of 245 (standard: 116; early fast-track: 129) included in the analyses. The final study visit occurred on September 6th, 2023.

**Fig 1.**
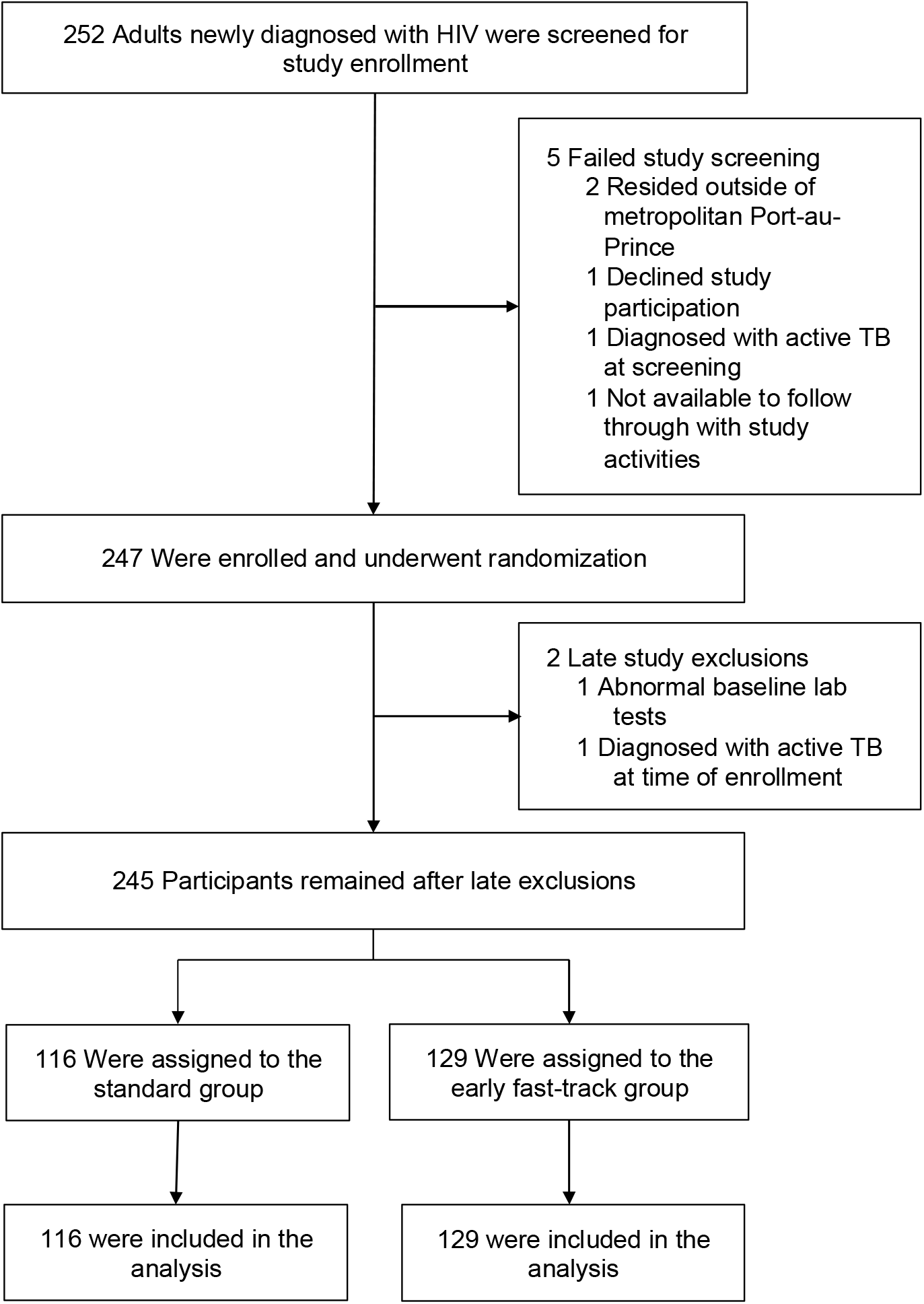
CONSORT Diagram: Screening, Randomization, and Analysis Populations.

Demographic and clinical characteristics are described in **Table 1**. Median time from HIV diagnosis to study enrollment was 0 days (interquartile range [IQR]: 0, 0). Among the 116 participants in the standard group, 81 (69.8%) received 24-week viral load testing, and 73 (90.1% of those tested; 62.9% of those randomized) had 24-week <200 copies/mL and were eligible for fast-track care. Among the 129 participants in the early fast-track group, 100 (77.5%) received 8-week viral load testing and 97 (97.0% of those tested; 75.2% of those randomized) had HIV-1 RNA <200 copies/mL. Of the remaining 32 participants in the early fast-track group (with missing or elevated 8-week viral load), 15 received 12-week viral load, and 11 (73.3%) had HIV-1 RNA <200 copies/mL. In total, 108 (83.7%) of participants in the intervention group initiated fast-track care within 12 weeks after enrollment.

**Table 1.** Baseline Characteristics of Study Participants by Group.

|  | Standard Care Group<br>(n=116) | Early Fast-Track Group<br>(n=129) |
| --- | --- | --- |
| Female – no. (%) | 46 (39.7) | 64 (49.6) |
| Age at enrollment (years) – median (IQR) | 40 (36, 47) | 40 (33, 48) |
| Income – no. (%) |  |  |
| <\$US 1.00 per day | 82 (70.7) | 93 (72.1) |
| ≥\$US 1.00 per day | 14 (12.1) | 10 (7.8) |
| Missing | 20 (17.2) | 26 (20.2) |
| Education level – no. (%) |  |  |
| None | 16 (13.8) | 11 (8.5) |
| Preschool or some primary | 28 (24.1) | 38 (29.5) |
| At least some secondary | 52 (44.8) | 54 (41.9) |
| Missing | 20 (17.2) | 26 (20.2) |
| Marital Status – N (%) |  |  |
| Single | 29 (25.0) | 30 (23.3) |
| Married | 46 (39.7) | 42 (32.6) |
| Formerly married | 21 (18.1) | 31 (24.0) |
| Missing | 20 (17.2) | 26 (20.2) |
| CD4 count (cells/mm<sup>3</sup>)—median (IQR) | 293 (154, 451) | 318 (191, 485) |
| CD4 count category—N (%) |  |  |
| <100 cells/mm<sup>3</sup> | 14 (12.1) | 9 (7.0) |
| 100-349 cells/mm<sup>3</sup> | 43 (37.1) | 39 (30.2) |
| 350-499 cells/mm<sup>3</sup> | 17 (14.7) | 15 (11.6) |
| ≥500 cells/mm<sup>3</sup> | 19 (16.4) | 19 (14.7) |
| Missing | 23 (19.8) | 47 (36.4) |

Among the 116 participants in the standard group, 106 (91.4%) were retained, and 78 (67.2%) participants received 48-week viral load testing (**Table 2**). Sixty-six participants (84.6% of those tested; 56.9% of those randomized) had HIV-1 RNA <200 copies/mL. Among the 129 participants in the early fast-track group, 112 (86.8%) were retained and 87 (67.4%) received 48-week viral load testing. Seventy-nine (90.8% of those tested; 61.2% of those randomized) had HIV-1 RNA <200 copies/mL. There was no difference between groups (61.2% vs. 56.9%; RD: 0.043; 95% CI: -0.080, 0.167). There was also no difference between groups with HIV-1 RNA cut-offs of <50 copies and <1000 copies/mL.

**Table 2.** Primary and Secondary Outcomes by Group.

| **Outcome** | **Standard Care**<br>**Group (n=116)** | **Early Fast-Track**<br>**Group (n=129)** | **Risk Difference (95%**<br>**CI)** | **p-value** |
| --- | --- | --- | --- | --- |
| **Primary Outcome** |  |  |  |  |
| 48-week HIV-1 RNA <200 copies/mL | 66 (56.9%) | 79 (61.2%) | 0.043 (-0.080, 0.167) | 0.490 |
| Received 48-week viral load testing | 78 (67.2%) | 87 (67.4%) | 0.002 (-0.116, 0.120) | 0.973 |
| HIV-1 RNA <200 copies/mL among<br>those receiving viral load testing | 66/78 (84.6%) | 79/87 (90.8%) | 0.062 (-0.039, 0.162) | 0.224 |
| **Secondary Outcomes** |  |  |  |  |
| 48-week HIV-1 RNA <50 copies/mL | 61 (52.6%) | 74 (57.4%) | 0.048 (-0.077, 0.172) | 0.453 |
| 48-week HIV-1 RNA <1000 copies/mL | 70 (60.3%) | 82 (63.6%) | 0.032 (-0.090, 0.154) | 0.604 |
| *Forty-Eight Week Outcomes\** |  |  |  |  |
| Retained in care at 48 weeks | 106 (91.4%) | 112 (86.8%) | -0.046 (-0.123, 0.032) | 0.255 |
| Died | 2 (1.7%) | 1 (0.8%) | -0.009 (-0.038, 0.019) | 0.604 |
| Lost to follow up | 4 (3.4%) | 6 (4.7%) | 0.012 (-0.037, 0.061) | 0.752 |
| Fled home due to gang violence | 3 (2.6%) | 10 (7.8%) | 0.052 (-0.003, 0.106) | 0.072 |
| Transferred | 1 (0.9%) | 0 (0.0%) | -0.009 (-0.025, 0.008) | 0.473 |

\*These are mutually exclusive outcomes which include all 245 randomized participants.

In the standard group, 2 participants (1.7%) died, 4 (3.4%) were LTFU, 3 (2.6%) fled their homes due to gang violence, and 1 (0.9%) was transferred (**Table 2**). In the early fast-track group, 1 (0.8%) died, 6 (4.7%) were LTFU, and 10 (7.8%) fled their homes due to gang violence. There was no difference between groups in retention or mortality. A total of 60 (76.9%) in the standard and 65 (74.7%) in the early fast-track groups achieved adherence ≥90% (p=0.741). No participant stopped TLD due to adverse events. Participants in the standard care and early fast-track groups had a mean of 4.8 and 4.4 visits, respectively (MD: -0.308; 95% CI: -0.682, 0.066, p = 0.106).

## DISCUSSION

This study was conducted during the COVID-19 pandemic and a period of severe civil unrest and gang violence in Haiti, which presented extraordinary challenges to the health system. We found that early fast-track care (8 to 12 weeks after same-day TLD initiation) is associated with high rates of retention and viral suppression among patients with early-stage HIV disease. The findings of this study do not confirm our hypothesis that early fast-track care would improve viral suppression, compared to standard treatment. However, we note that early fast-track care offers the benefit of expedited visits and choice of visit location. It is noteworthy that though retention was high in both groups, one-third of patients were not able to receive 48-week viral load testing, due to lack of access to testing at community ART pick-up sites.

This study provides additional evidence in support of DSD models of care.^1-7,9,10^ GHESKIO’s fast-track intervention is part of a multicomponent strategy, which also includes same-day linkage to care, adherence counseling, and social support, which were also found to be effective in the SEARCH trial in Kenya and Uganda.^14,15^ In addition, patients receive same-day ART initiation, with an effort by staff to make them feel welcome, and avoid unnecessary delays in care, which have also been found to be important in other studies.^16-23^ After ART initiation, patients are provided with decentralized care and multi-month prescribing, which have been widely implemented in LMICs.^6,24-26^

Results in both groups in our study were outstanding, in spite of the COVID-19 pandemic and severe civil unrest in Haiti. At 48 weeks after HIV diagnosis, approximately 90% in both groups were retained in ART care; among those tested, viral suppression rates were 85% in the standard and 91% in the early fast-track group. These outcomes equal or surpass the intervention groups in other same-day ART studies, including those conducted at our site.^16-18,20,27^ We attribute this to the use of the high potency and tolerability of DTG, as well as streamlined provision of follow-up care. GHESKIO staff are also encouraged to create a welcoming environment and trained in strategies to reduce stigma and support patient adherence; these interventions have also been incorporated into other successful rapid ART programs.^16-18,20,21,28,29^

Since completion of this study, viral load testing capacity has been expanded at the community sites in GHESKIO’s network. Point-of-care HIV-1 RNA tests, which are currently in development, could also greatly facilitate viral load testing in this context. We note that other DSD models of care have also been found to be effective in settings of civil unrest.^30^ A study of PLWH with stable disease in the Democratic Republic of the Congo also documented a high rate of retention at community-based ART centers, though patients were classified as stable after at least six months on ART.^29^ A project conducted in a conflict-affected area of South Sudan, which included mobile teams with HIV testing and same-day ART initiation, documented high rates of retention and viral suppression.^31^ In the Central African Republic and Yemen, contingency plans to distribute ART, including multi-month ART dispensing and health information cards, were successfully implemented during periods of acute conflict.^32^

Our study was conducted among ART-naïve patients in a large urban clinic, which may limit the generalizability of our findings. However, we note that many components of the GHESKIO HIV care model are also widely implemented through PEPFAR programs in other settings.

## CONCLUSIONS

In conclusion, our results indicate that providing fast-track care at eight to 12 weeks after TLD initiation appears to result in high levels of retention in care and viral suppression in a setting of severe civil unrest and gang-related violence. Further research is indicated to determine if earlier transition to DSD models of care may also be effective in other settings. Completion of 48-week viral load testing was suboptimal; point-of-care testing capacity may facilitate completion of viral load testing in this setting.

## Data Availability

All data produced in the present study are available upon reasonable request to the authors.

## Acknowledgments

We thank the patients who participated in this study and the GHESKIO staff who cared for them. We also thank Vince Marconi, Jonathan Colasanti, Robert Peck, and Margaret McNairy of the Data Safety Monitoring Board, the Community Advisory Board, and the ethics committees for their expertise and advice. This study was funded by a grant from ViiV Healthcare.

## Author Contributions

All authors were involved in drafting the article, and all read and approved the final version to be published. Study conception and design: SP, PS, CG, JWP, and SPK: Acquisition of data: JBM, SP, OD, FH, VRR, ND, CG, VR, CFD, GPLF, JJ, GB, EDD, and RCBR; Analysis and interpretation of data: JBM, SP, FH, AZ, VRR, PS, VR, ITK, JWP, and SPK.

